# Digital PCR Linkage Analysis Resolves *Streptococcus pneumoniae* Signature from Commensal Interference in Saliva Samples: Identifying Wolves among Sheep in Wolf’s Clothing

**DOI:** 10.1101/2025.08.02.25332851

**Authors:** Willem R. Miellet, Tessa Nieuwenhuijsen, Eline H.M.R. van den Oetelaar, Janieke van Veldhuizen, Nynke Y. Rots, Krzysztof Trzcinski, Rob Mariman

## Abstract

**Background:** Serotyping of *Streptococcus pneumoniae* (pneumococcus) is essential for evaluating the effects of conjugate polysaccharide vaccines on asymptomatic colonization (carriage) of vaccine-targeted capsular variants (serotypes). However, culture-independent application of molecular methods is hampered by genetic exchange between pneumococcus and oral streptococci co-residing within the airways. Unlike qPCR, digital PCR (dPCR) enables linkage analysis that allows for the identification of gene co-occurrence within individual bacterial cells. To minimize false-positive classification with molecular serotyping of highly polymicrobial saliva samples, a duplexed dPCR protocol for linkage analysis was developed.

**Methods:** Performance of the protocol was evaluated by determining linkage between *piaB* and *lytA,* as well as between *piaB* and genes coding for serogroups 6, 9 and serotype 4 capsular polysaccharides in saliva samples from children and adults.

**Results:** Identification of linkage between distant genes required intact pneumococcal cells. The 95% limit of detection for linkage analysis between *piaB and lytA* was determined to be 9.2 CFU/reaction. Co-occurrence of *piaB* and *lytA* within single bacterial cells was consistently observed in saliva samples. Similarly, linkage was identified between serogroup 6 and *piaB* in the majority of saliva samples from serogroup 6 carriers. Application of dPCR distinguished pneumococcus from oral streptococcus species in saliva tested for serogroup 9 and serotype 4.

**Discussion:** Duplex dPCR linkage analysis allowed for differentiation of pneumococcus from oral streptococci with homologous capsular loci. By improving the diagnostic accuracy of molecular surveillance, linkage analysis is a promising technique for culture-independent epidemiologic surveillance of pneumococcal serotype carriage.

## INTRODUCTION

*Streptococcus pneumoniae* (pneumococcus) is a commensal bacterium of the human upper respiratory tract (URT) but also a major cause of disease, with the highest incidence in young children and older adults. The polysaccharide capsule is considered to be the primary virulence factor of pneumococcus. Over 100 variants (serotypes) of *S. pneumoniae* have been described, classified based on capsular polysaccharide immunogenicity [1]. Capsular polysaccharides are also the antigens in all currently marketed pneumococcal vaccines [1]. Most broadly used vaccines are pneumococcal conjugate vaccines (PCVs) that prevent not only disease but also acquisition of carriage with vaccine-targeted (VT) serotypes [2]. Consequently, carriage of these serotypes is accepted as an endpoint in vaccine impact studies [3].

Among young children, colonization rates of both the nasopharyngeal and oral mucosa are high [4–7]. Due to high absolute and relative abundances of *S. pneumoniae* within nasopharynx, it is relatively straightforward to detect pneumococcal carriage through culture of nasopharyngeal samples in this age group. In contrast, culture-based methods have proven inadequate for the sensitive detection of *S. pneumoniae* at highly polymicrobial sites such as the oropharyngeal and oral mucosa [8]. This is particularly problematic in adults, among whom pneumococcal colonization of upper respiratory tract is largely restricted to oropharynx and oral cavity [9]. Disparity between culture and culture-independent methods for such samples mirrors the ‘great plate count anomaly’ due to the inadequacy of culture-based methods to highly polymicrobial samples [10]. Consequently, oral and oropharyngeal samples are often overlooked for pneumococcal carriage detection.

Molecular diagnostic methods can be used to accurately identify *S. pneumoniae* and pneumococcal serotypes carriage in individuals across various age groups [8]. However, the utility of molecular diagnostic assays to oral samples is at times complicated by habitat overlap of *S. pneumoniae* and other oral streptococci (*S. mitis* and *S. oralis*, *etc*.) and frequent genetic exchange between these bacteria [11, 12]. Although recent advances in qPCR-based carriage surveillance have enabled sensitive evaluation of diagnostic accuracy through the application of the Two-to-Tango approach [8, 13], interspecific horizontal gene transfer has rendered numerous qPCR assays unreliable in multiple geographic settings, including assays designed to detect serotypes that are targeted by licensed PCVs [8]. Like sheep in wolves’ clothing, the circulation of such non-pneumococcal streptococci with homologous capsular operons has complicated the interpretation of molecular serotyping results. Consequently, while we and others have defined certain limitations of culture-independent surveillance methods [8, 14, 15], some of our colleagues have altogether discouraged the application of carriage surveillance on oral samples [1, 16], and culture-independent testing of oral fluids is currently not recommended by the World Health Organization Pneumococcal Carriage Working Group [17].

Here, we describe a digital PCR (dPCR) based protocol capable of confirming *S. pneumoniae* as the source of serotype signature in polymicrobial samples. By partitioning duplexed molecular diagnostic tests into many micro reactions, it becomes possible to identify whether two targets are non-randomly linked, either physically on the bacterial genome or as part of the same bacterial cell (*e.g*., bacterial chromosome and plasmid). While unlinked targets distribute randomly across partitions, linked targets tend to co-localize consistently in the same partitions, resulting in a high ratio of double-positive to single-positive partitions. By using intact bacterial cells as input for dPCR [18, 19], we assessed linkage between physically distant targets to address specificity limitations of various serotype/serogroup-specific PCR assays, namely serotype 4, serogroup 6 (6A/B/C/D), and serogroup 9 (9A/L/N/V). We validated this approach by using suspensions of mixed cells of defined streptococcal strains. Finally, we demonstrated the utility of the protocol by applying it to culture-enriched saliva samples collected from community-dwelling individuals.

### EXPERIMENTAL MODEL AND SUBJECT DETAILS

#### Microbial strains

All bacterial strains used in this study are listed in **Table S1**. A non-pneumococcal Streptococcus isolate cultured from the upper airways in the carriage study and found by qPCR to be positive for serogroup 9 capsular operon (*cps*) isolate, yet negative for *piaB* and *lytA* (strain Sm226001019702), was used alongside *S. pneumoniae* strain P2007-1850 (serotype 9A) to evaluate the performance of duplex digital PCR (dPCR) in detecting gene linkage in mixed streptococcal samples. For comparison of DNA extract versus intact cell input and as a positive control in *piaB-lytA* duplex assays, the *S. pneumoniae* strain Hungary 19A-6 (serotype 19A) was used. Strains PI2018-0103 (6A), PI2013-2681 (6C), P2007-1850 (9A), and PI2018-0984 (4) were included as serotype-specific positive controls in dPCR analyses targeting each respective serotype together with *piaB*.

#### Human subjects

Pneumococcal carriage was investigated in cross-sectional prospective observational study conducted in 2015/2016 in the Netherlands [20]. The study was approved by the Medical Ethics Committee Noord Holland (NL53027.094.15 on onderzoekmetmensen.nl/nl/trial/42659). Written informed consent was obtained from the parent or guardian of every participating child, and adults produced written consent for their own participation. The study was conducted in accordance with the Declaration of Helsinki and Good Clinical Practice.

## METHOD DETAILS

### Sample collection and laboratory processing

As described previously [21]. In short, saliva samples were collected from all individuals with sponge lollipop (Oracol Saliva Collection System Malvern Medical Developments Limited, Worcester, UK), immediately transferred to tubes pre-filled with 100 μl sterile 50% glycerol solution in water, mixed, placed on dry ice and transported to the lab. With approximately 400 μl of saliva collected per sample the final glycerol concentration was around 10%. Saliva samples were delivered to the laboratory and stored at −70°C within 8 hours. For dPCR analysis, saliva samples from children and adults were included if they were previously identified as positive for both *piaB* and *lytA*, and, in cases involving molecular serotyping, also positive for the relevant serotype assay [5]. In addition, selected samples were chosen to span a C_q_ range representative of the full sample set.

### Study sample culture-enrichment

Glycerol-supplemented saliva was used to inoculate gentamicin sheep blood agar (SB7-Gent agar, Oxoid). After overnight incubation at 37°C and 5% CO_2_, all growth was harvested from a plate into 10% glycerol in brain heart infusion broth (BHI, Oxoid) supplemented with 0.5% yeast extract (Oxoid) [5]. Harvests from SB7-Gent plates were used to inoculate Columbia Blood Agar (CBA) with 5% defibrinated sheep blood and incubated for 6 hours at 37°C with 5% CO_2_. After incubation glycerol was added to a final concentration of 10% and harvests were stored at −70°C for further processing.

### Bacterial cell suspensions and CFU assessment

*S. pneumoniae* positive controls were prepared by growing the strains in BHI with 0.5% yeast extract to mid-logarithmic growth phase (OD₆₂₀_nm_ around 0.1) at 37°C with 5% CO_2_. The cultures were then placed on ice, and glycerol was added to a 10% final concentration before aliquoting and storing cell suspensions frozen at −70°C. Five eight-fold serial dilutions were prepared from an aliquot in phosphate buffered saline, the fourth and fifth dilutions were plated on CBA in duplicate. After overnight incubation at 37°C with 5% CO_2_, colony counts were used to calculate the concentration in CFU/ml. This information was used to assess the dynamic range of the dPCR linkage protocol and estimate the 95% limit of detection (LoD_95_).

### Primers and Probes

DNA was quantified in cell suspensions and DNA extracts with duplexed assays. Primers and probe targeting sequences within genes coding for the pneumococcal iron uptake ABC transporter lipoprotein PiaB [22], and for the major pneumococcal autolysin LytA [23]. were used to detect pneumococcal DNA. We used three assays for molecular serotyping in combination with *piaB*, namely assays targeting the serogroup 6ABCD (*wciP* gene), 9ALNV (*mnaA* gene) and serotype 4 (*wzy* gene) *cps* (**Table S2**). Primers and probes used in these assays, and their concentrations are also listed in **Table S2**.

### Nucleic acid extraction & quantitative PCR (qPCR)

As described previously [5]. DNA was extracted from culture-enriched saliva by centrifuging 100 μl of plate harvest (2 min, 14,000 × g), resuspending the pellet in 90 μl TE buffer (20 mM Tris-HCl, 2 mM EDTA, pH 8.0), and incubating at 95°C for 15 min. After adding 90 μl lysis buffer (20 mM Tris-HCl, 2 mM EDTA, 2.4% Triton X-100, 40 mg/ml lysozyme), DNA was purified using the DNeasy Blood & Tissue Kit (Qiagen) and eluted in 200 μl buffer. Next, 1.0 µl DNA extract within a total reaction volume of 12.5 µl with SensiFast Probe No-Rox mastermix (BIO-86050, Bioline) was tested with a LightCycler480 qPCR unit (Roche). The thermocycler conditions were as follows: one cycle of 95°C for 10 minutes, followed by 45 cycles of 95°C for 10 seconds and 60°C for 45 seconds. C_q_ calling was performing according to manufacturer instructions with a standard curve and via the second derivative maximum method.

### Digital PCR (dPCR)

dPCR was performed using the QIAcuity One 2plex dPCR instrument (cat.nr. 911000, Qiagen) according to the manufacturer’s recommendations. Reactions of 40 µl were prepared with QIAcuity Probe mastermix (cat.nr. 250101, Qiagen) and loaded in a QIAcuity 26k 24-well plate (cat.nr. 250001, Qiagen) in partition volumes of approximately 0.82 nl. Unless stated otherwise, the thermocycler conditions were as follows: QIAGEN Priming Profile Probe (RT-) PCR, one cycle of 95°C for 5 minutes, followed by 55 cycles of 95°C for 30 seconds and 60°C for 60 seconds. The imaging settings of duplex reactions used for linkage analysis are listed in **Table S2**. All plates included at least one no-template-control and positive control (either DNA extract or log-phase cell suspension of a pneumococcal strain). Study samples were diluted prior to testing based on prior qPCR results: samples with *piaB*/serotype C_q_ values <22 were diluted 1:1000, those with C_q_ values between 22 and 25 were diluted 1:100, and those with C_q_ ≥25 were diluted 1:10. All samples were tested in singlicate.

### Baseline correction and RFU Thresholding

Relative fluorescence unit (RFU) thresholds were derived using the method described by Trypsteen *et al*. 2015 based on extreme value theory [24]. Prior to RFU thresholding, baseline correction was conducted as outlined in the same study [24]. In short, all no template controls (NTCs) measurements from a particular duplexed assay were combined and split into blocks of 150 partitions. The initial thresholds were set to the 0.9995 quantile, and the procedure was iterated 10,000 times. Estimated RFU thresholds were thereafter manually curated and in case of inaccurate RFU thresholding initial thresholds and block size were adjusted. Details related to the Minimal Information for MIQE Experiments (dMIQE) guidelines for digital droplet PCR reporting are outlined in **Table S4** [25].

### Linkage analysis

Concentration-dependent linkage (linkage for short) was calculated according to method described by Regan *et al*. 2015 [26]. As these values are inherently influenced by target concentration and given the typically low abundance of *S. pneumoniae* in oral fluids, concentration-independent linkage was also calculated for each target within a duplex reaction using the following approach. If two targets are unlinked their distribution among partitions follows the Poisson distribution:

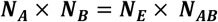

Where ***N*** indicates the number of partitions analyzed with ***N_A_*** and ***N_B_*** the number of partitions positive for a single target, ***N_AB_***the number of double-positive partitions and ***N_E_*** the number of double-negative partitions. By rearranging the equation, we can then calculate the number of double-positive partitions by random chance (***N_Ch_***):

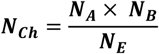

When linkage is observed within a reaction, *N_AB_ > N_Ch_*. For such samples the number of partitions attributable to linkage (***N_L_***) can be calculated with:

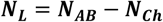

Using this information we can calculate concentration-independent linkage values via the following formulas:

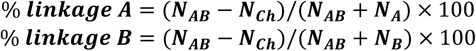

Concentration-independent linkage values were subsequently used for all analyses. Unless stated otherwise, linkage values are reported with *piaB* as the reference target. A sample was classified as positive for linkage when ***N_L_*** was ≥5 and a two-tailed Poisson test (with α=0.01) indicated linkage, meaning that the 99% confidence interval of ***N_AB_*** did not include ***N_Ch_***.

### Bland-Altman analysis

Presence or absence of linkage in relation to mean target concentration and target agreement was illustrated using Bland-Altman plots. Non-parametric limits of agreements were used to visualize the degree of agreement in dPCR measurements between duplexed targets, to assess potential bias, defined as the ratio of copies per microliter (cp/µl) of assay A to that of assay B [27, 28].

### Probit regression analysis

The LoD_95_ was derived by fitting a probit regression model to the counts of successful and failed detections of linkage at specific CFU/ml concentrations. This was done using the *glm* function from the ‘stats’ R package, with a binomial family and probit link. All statistical analyses were performed in R v4.4.3 using Rstudio (2024.12.1-563).

## RESULTS

### Pneumococcal Cell Suspensions, but not DNA Extracts, display Linkage between Two Targets

Using dPCR, we subjected suspensions of intact cells and DNA extracts of *S. pneumoniae* Hungary 19A-6 strain to duplex quantification to assess the linkage between two genes, *piaB* and *lytA*, a combination of targets presumed to be highly specific for pneumococcus [29]. In both types of samples, quantification of both molecular targets was observed (**Fig. 1B**). Suspensions with intact pneumococcal cells consistently produced partitioned reactions with co-amplification of *piaB* and *lytA* beyond a number expected due to chance (**Fig. 1A** and **C**). By contrast, duplex dPCR reactions with DNA extracts exhibited no linkage between the two targets (**Fig. 1A** and **1D**). The number of partitions with co-amplification of *piaB* and *lytA* in these reactions corresponded with the number expected due to chance (**Fig. 1D**). Such results were interpreted as evidence of linkage between two genes, meaning both genes were derived from a single bacterial cell. Absence of linkage with DNA extracts was hypothesized to be due to the considerable distance between *piaB* and *lytA* in the typical pneumococcal genome. In the *S. pneumoniae* Hungary 19A-6 strain these two genes are approximately 876 kb apart, a distance exceeding the maximum fragment size of genomic DNA extracts that are typically ∼30.0000 bp (**Fig. 1E**).

**Figure 1:**
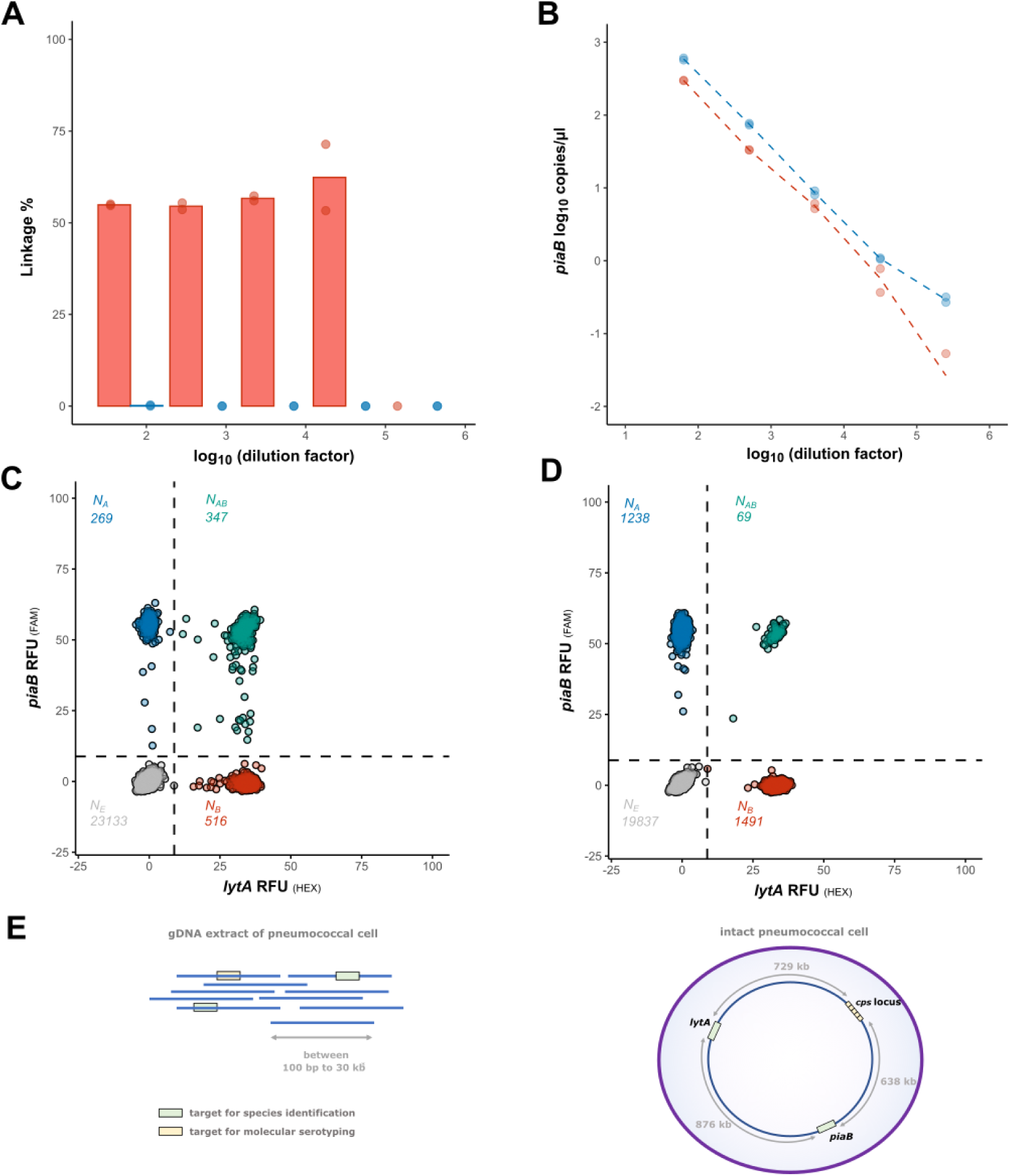
Linkage between *piaB* and *lytA* for intact cells and DNA extracts of *S. pneumoniae* Hungary 19A-6 strain. **A**: Linkage detection between genetic loci relies on either the genomic distance or the physical proximity of targets. If targets are separated by more than the average DNA fragment length, co-amplification becomes stochastic. **B**: Utilizing intact bacterial cells as input material permits the identification of linkage between targets that are genomically distant. Linkage between distant targets is lost when DNA extracts are used. **C:** Quantification of *piaB* with intact cells (red) and DNA extracts (blue) at a given dilution of an input sample. Bacterial cell suspensions were 1.87E^2^ to 6.12E^6^ CFU/ml and DNA extracts were 3.5E-2 to 1.1E-6 ng/µl, equivalent of 0.3 to 793 cp/µl. Linkage % estimates were calculated with *piaB*-positive partitions as the denominator. **C** Linkage observed with intact cells (red) and DNA extracts (blue) at a given concentration. **D**: Representative 2D plot of bacterial cell suspension of 14 cp/µl (*piaB*), the ratio of double-positive to single-positive partitions is suggestive of linkage (347 × 23133 > 269 × 516). **E**: Representative 2D plot of DNA extract of 32 cp/µl (*piaB*). The ratio of double-positive to single-positive partitions was not indicative of linkage (69 × 19837 ≈ 1238 × 1491).

### Critical Determinants for Linkage Analysis

It was reported that raising the number of PCR cycles, pre-treatment bacterial cells with lysozyme, or supplementing reaction mixtures with bovine serum albumin (BSA) concentration may lead to an increase in percentage linkage or improved PCR performance [30–32]. The impact of PCR cycle number on linkage was tested by a stepwise increase by 5 cycles from 40 recommended by the manufacturer to 60 (data not shown). Raising the number of cycles to 55 was associated with a modest increase in percentage linkage. Neither treatment of pneumococcal cells with lysozyme, nor increasing the concentration of BSA resulted in an increase in linkage (data not shown). Of note, cell suspensions were tested exclusively on 26k well plates, as the 8.5k variant was prone to clogging thus rendered unsuitable for dPCR with whole bacterial cells (data not shown).

### Analytical Sensitivity Estimation *via* Probit Regression Analysis

The analytical sensitivity of the protocol for detecting linkage was evaluated using probit regression with a 2-fold serial dilution of *S. pneumoniae* Hungary 19A-6 strain cells, ranging between 1.20E+04 to 1.87E+02 CFU/ml, with twelve replicates tested for each dilution. LoD_95_ was estimated to be 1664 CFU/ml, which corresponded to 9.2 CFUs per reaction or 0.8 copies/µl (**Fig. S1**). Importantly, to minimize false-positive linkage detection in samples with faint PCR target quantification, a sample was deemed positive for linkage when a minimum of five partitions showed co-amplification not attributable to chance (N_L_) and a Poisson test of observed (N_AB_) versus expected partitions with co-amplification (N_Ch_) indicated linkage (N_AB_>N_Ch_).

### In Mixtures of Streptococcal Strains, dPCR-based Linkage is Observed only if *S. pneumoniae* is Present

To emulate the complexity of polymicrobial samples with mixtures of pneumococci and oral streptococci, we evaluated performance of the protocol applied to bacterial cell suspensions with varying ratios of serotype 9A *S. pneumoniae* strain P2007-1850 (**Fig. 2A**) to a non-pneumococcal oral streptococcus strain Sm2260010119702 (**Fig. 2B**). Both strains were positive by qPCR for *mnaA* gene unique for serogroup 9, therein further described as serogroup 9 *cps* assay, yet only the *S. pneumoniae* was positive for *piaB* (**Table S2**). Linkage between *piaB* and serogroup 9 *cps* was only detectable in mixes containing *S. pneumoniae* cells provided the pneumococcal abundance was above the LoD_95_ (**Fig. 2D**). Linkage remained detectable in mixes where the non-pneumococcal strain exceeded pneumococcus by up to 100-fold (**Fig. 2C**). In these mixes, the percentage of linkage estimated by *piaB* remained consistent, while estimated by the serogroup 9 *cps* target decreased with increasing relative abundance of the non-pneumococcal strain (**Fig. 2C** and **D**). Taken together, linkage remained identifiable in mixes, suggesting the potential value of the protocol to complex polymicrobial samples such as saliva.

**Figure 2:**
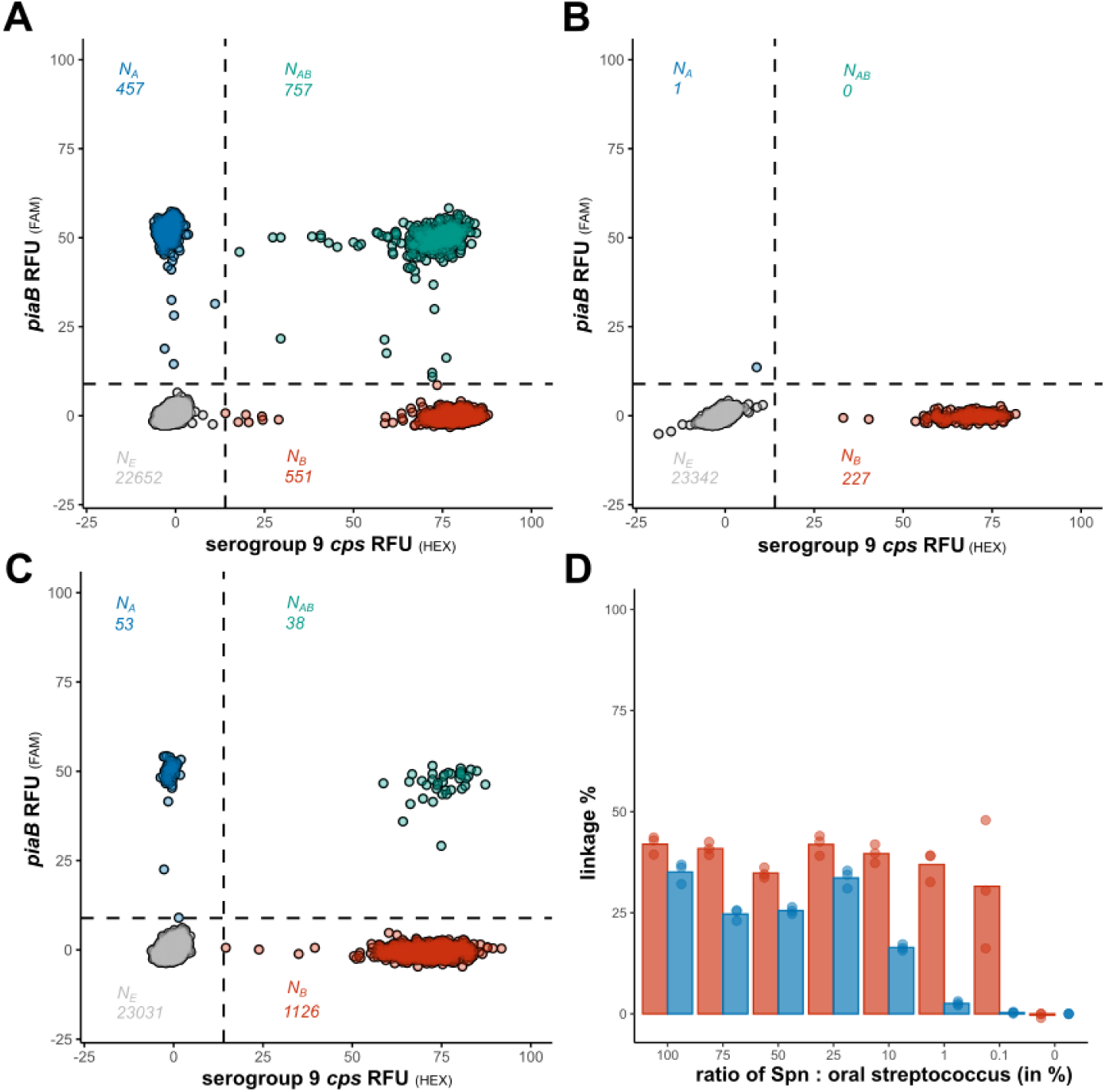
dPCR resolves the *S. pneumoniae* serogroup 9 *cps* signature in mixed suspensions containing both pneumococcal and non-pneumococcal strains. A: A 2D plot of the serogroup 9 *cps S. pneumoniae* strain with linkage evident between *piaB* and the serogroup 9 *cps* assay. **B**: A 2D plot of the serogroup 9 *cps* non-pneumococcal strain, which only produced quantification for the serogroup 9 *cps* assay and not for *piaB*, hence no linkage was evident. **C**: A 2D plot of a suspension with 1:100 ratio of serogroup 9 *cps S. pneumoniae* strain and non-pneumococcal serogroup 9 *cps* strain, linkage between *piaB* and serogroup 9 *cps* assay remains identifiable. **D**: Bar diagram of percentage linkage of suspensions of serogroup 9 *cps S. pneumoniae* strain mixed with non-pneumococcal serogroup 9 strain in various ratios. Red and blue bars indicate linkage percentage based on *piaB* and serogroup 9 *cps* assay, respectively.

### Assessment of Linkage in Saliva Study Samples and Optimization of Culture Conditions

In the experiments that followed saliva samples collected from community-dwelling individuals were tested dPCR for linkage between pneumococcal genes. All these samples were previously culture-enriched for pneumococcus and tested for *S. pneumoniae* and pneumococcal serotypes using a qPCR-based protocol [5]. On top of that, paired nasopharyngeal samples collected from the same individuals were tested using WHO recommended method of cultured pneumococcal isolate serotyping of with Quellung. When samples of culture-enriched saliva were tested in duplex dPCR targeting *lytA* and *piaB* percentage linkage detected was lower than expected based on tests with pneumococcal cell suspensions (**Fig. S2A** and **B**). It implied that the number of viable pneumococcal cells plummeted in these samples due to either fratricide or bactericidal effects of co-cultured bacteria in saliva cultures incubated in the original study for a relatively long period of overnight [33]. To test for this, we compared different culture incubation times for *S. pneumoniae* Hungary 19A-6 strain. Linkage percentage was observed to be highest after six hours (**Fig**. **S2C** and **D**). Accordingly, fresh 6-hour cultures were prepared from n=43 study samples of culture-enriched saliva previously identified as positive for *S. pneumoniae* by qPCR (**Fig**. **3A** and **B**), with C_q_ values ranging from 20 to 36 C_q_ (among original samples), or equivalent of 250 to 0.05 cp/µl. Linkage between *piaB* and *lytA* was observed in 26 of 43 (60.5%) 6-hour culture samples (**Fig. 3C**), with linkage percentages ranging from 13.3% to 83.3% (mean 34.9%; **Table S3**), and with samples that tested negative for linkage typically displaying quantification of *piaB* and or *lytA* below the LoD_95_.

**Figure 3:**
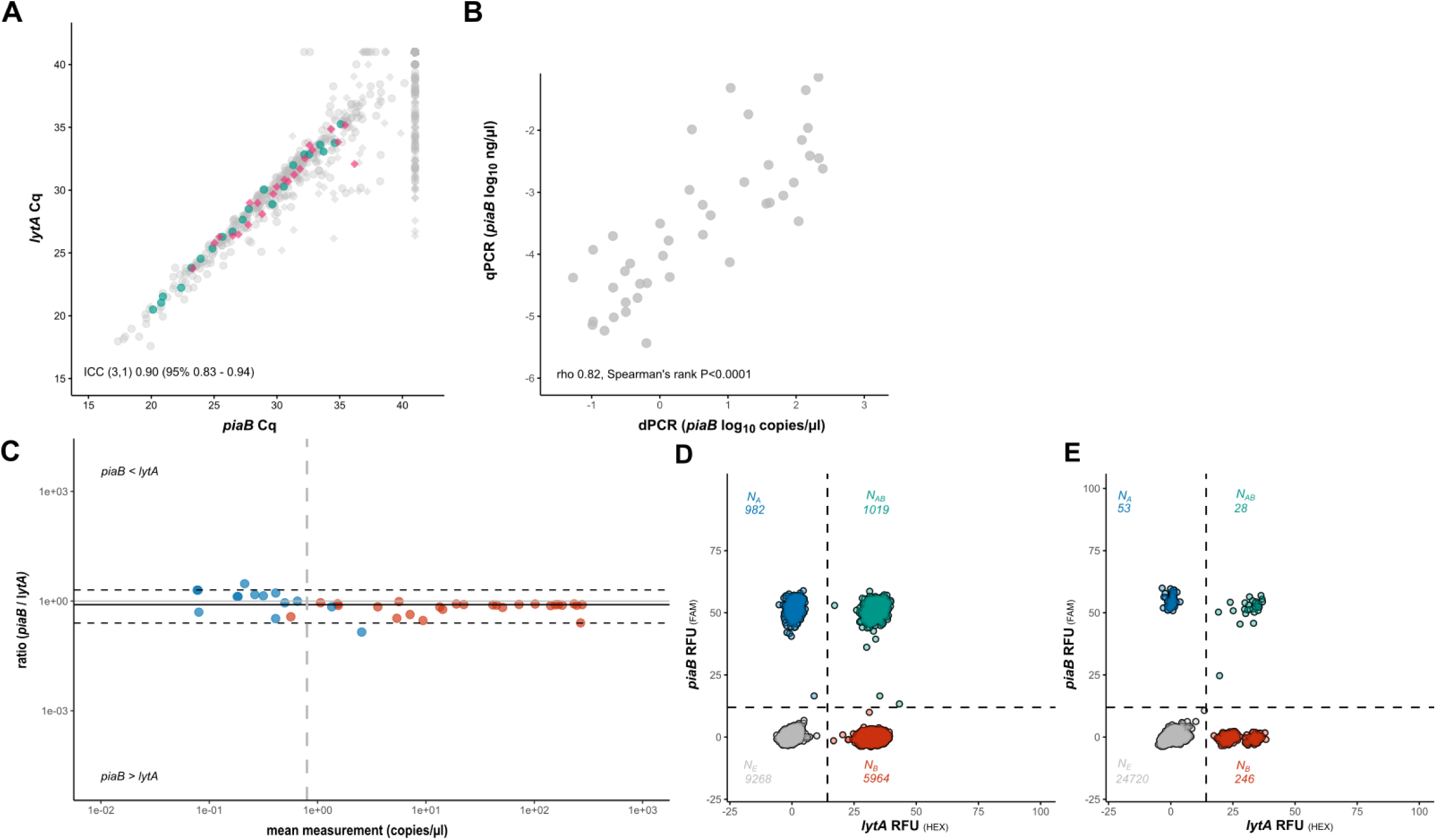
dPCR-based linkage analysis provides evidence for positive conditional dependence of *piaB* with *lytA* signal in culture-enriched saliva samples, which is associated with *S. pneumoniae* presence. A: qPCR-based quantification (in C_q_) of *piaB* and *lytA* in DNA extracts of culture-enriched saliva study samples. Samples from children and adults that were selected for dPCR-based linkage analysis are highlighted with green and pink respectively. The remaining samples are colored grey. Circles and diamonds indicate samples from children and adults, respectively. **B:** The correlation between cultures suggests that pneumococcal presence, as measured by *piaB*, was retained in saliva cultures after preparation of 6-hour cultures (dPCR) from the original overnight cultures (qPCR). **C**: non-parametric Bland-Altman plot of dPCR-based quantification (in copies/µl) *piaB* and *lytA* in culture-enriched cell suspensions, samples positive for linkage between *piaB* and *lytA* are highlighted with red and negative samples with blue. The vertical line indicates the LoD_95_, dashed horizontal lines the limits of agreement and solid line the bias (median difference between paired *piaB* and *lytA* measurements). **D**: An example of a 2D dPCR plot wherein linkage a relatively high number of partitions with co-amplification of *piaB* and *lytA* is observed, a result suggestive of linkage. **E**: An example of a 2D dPCR plot showcasing linkage between *piaB* and *lytA* and concurrent presence of oral streptococcus (or non-typeable pneumococcus) which is exclusively positive for *lytA*.

### dPCR Allows for Molecular Serotyping of Saliva Study Samples Positive for Pneumococcus

Six-hour cultures were prepared from a subset of n=43 saliva study samples that had previously tested positive by qPCR (**Fig. 4A**) pneumococcal molecular targets and, in assay targeting *wciP* gene, as positive for serogroup 6 *cps*. These include n=22 saliva samples from individuals (21 children and one adult) positive for serogroup 6 *S. pneumoniae* by WHO-recommended conventional culture method applied nasopharyngeal swab. Linkage between *piaB* and the serogroup 6 *cps* was detected, as long as the quantification of one of the two targets was not below the LoD_95_ (**Fig. 4C**). In total 21 of 43 (48.8%) samples were deemed positive for serogroup 6 pneumococci based on detected linkage estimated by *piaB*. Fourteen (57.1% of 21) linkage positive saliva samples were from individuals that were also positive for serogroup 6 pneumococci by nasopharyngeal culture. Among the five samples negative for linkage yet positive for quantification of the LoD_95_ for both targets, one sample exhibited RFU amplitude indicative of serotype 6A (data not shown), suggesting this sample may harbor a *piaB*-deficient *S. pneumoniae* strain, although an oral streptococcus with serogroup 6 *cps* could not be excluded.

**Figure 4:**
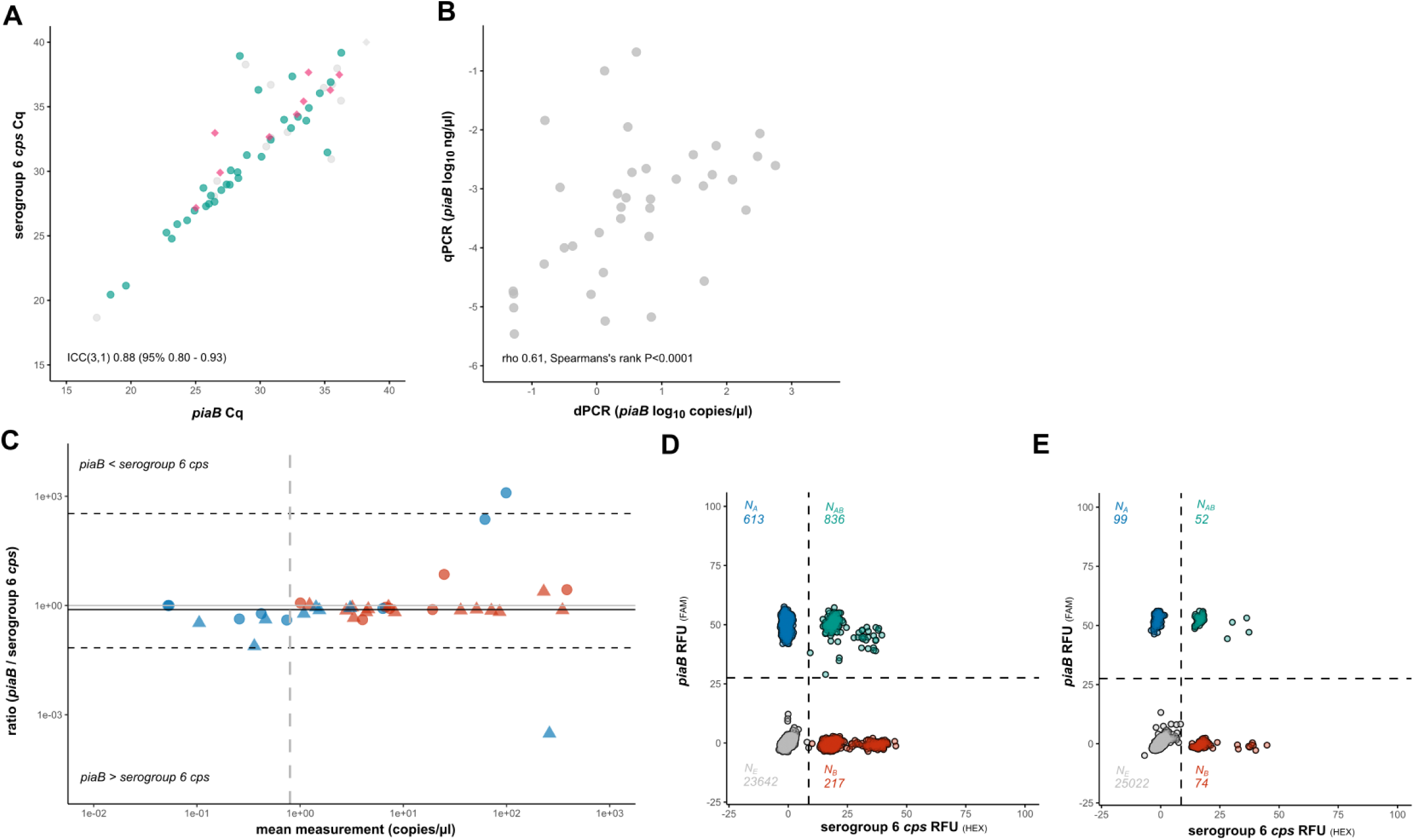
dPCR-based linkage analysis enables accurate molecular detection of serogroup 6 *S. pneumoniae* in culture-enriched saliva samples. A: qPCR-based quantification (in C_q_) of *piaB* and serogroup 6 *cps* in DNA extracts of culture-enriched saliva study samples. Samples from children and adults that were selected for dPCR-based linkage analysis are highlighted with green and pink respectively. The remaining samples are colored grey. **B:** The correlation between cultures suggests that pneumococcal presence, as measured by *piaB*, was retained in saliva cultures after preparation of 6-hour cultures (dPCR) from the original overnight cultures (qPCR). **C**: non-parametric Bland-Altman plot of dPCR-based quantification (in copies/µl) *piaB* and serogroup 6 *cps* in culture-enriched cell suspensions, samples positive for linkage between *piaB* and serogroup 6 *cps* are highlighted with red and negative samples with blue. Circles and triangles indicate samples from culture-negative and culture-positive individuals, respectively. Vertical line indicates the LoD_95_, dashed horizontal lines the limits of agreement and solid line the bias (median difference between paired *piaB* and serogroup 6 *cps* measurements). **D:** An example of a 2D dPCR plot wherein linkage is observed between *piaB* and serogroup 6 *cps*. An example of a high abundance sample. **E:** An example of a 2D dPCR plot wherein linkage is observed between *piaB* and serogroup 6 *cps*. An example of a low abundance sample.

### dPCR Linkage Analysis Resolves Pneumococcal Serotype Signature from Commensal Interference

Next, linkage analysis between *piaB* and serogroup 9 *cps,* was assessed for a subset of 43 saliva study samples from n=29 children and n=13 parents that were previously identified as positive for both targets by qPCR (**Fig. 5A**). The set included samples from four individuals (three children and one adult) whose paired nasopharyngeal samples were culture-positive for serotype 9N *S. pneumoniae*. Linkage was detected in four samples (**Fig. 5C**), all from abovementioned individuals from whom serotype 9N pneumococci were cultured. Of the remaining 39 samples negative for linkage between *piaB* and serogroup 9 *cps* seven were positive for both targets above the LoD_95_. Since the serogroup 9 *cps* assay was among assays classified as unreliable in oral samples, these seven samples likely contained oral streptococci harboring the sequence targeted in serogroup 9 *cps* assay.

**Figure 5:**
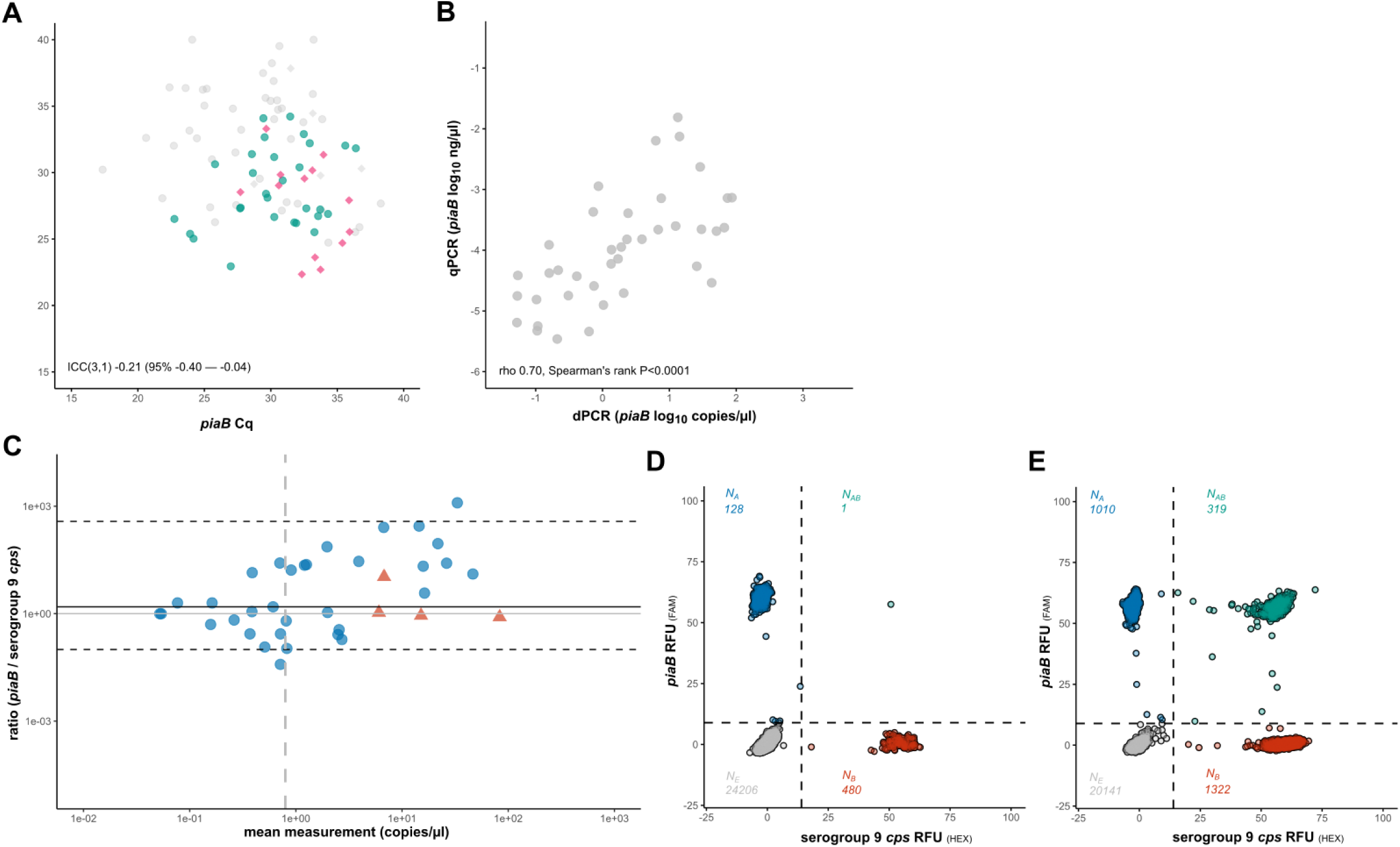
dPCR-based linkage analysis can distinguish between serogroup 9 *S. pneumoniae* and serogroup 9 signal from oral streptococci. A: qPCR-based quantification (in C_q_) of *piaB* and serogroup 9 *cps* in DNA extracts of culture-enriched saliva study samples. Samples from children and adults that were selected for dPCR-based linkage analysis are highlighted with green and pink, respectively. The remaining samples are colored grey. **B:** The correlation between cultures suggests that pneumococcal presence, as measured by *piaB*, was retained in saliva cultures after preparation of 6-hour cultures (dPCR) from the original overnight cultures (qPCR). **C:** non-parametric Bland-Altman plot of dPCR-based quantification (in copies/µl) *piaB* and serogroup 9 *cps* in culture-enriched cell suspensions, samples positive for linkage between *piaB* and serogroup 9 *cps* are highlighted with red and negative samples with blue. Circles and triangles indicate samples from culture-negative and culture-positive individuals, respectively. The vertical line indicates the LoD_95_, dashed horizontal lines the limits of agreement and solid line the bias (median difference between paired *piaB* and serogroup 9 *cps* measurements). **D:** An example of a 2D dPCR plot wherein no linkage is observed between *piaB* and serogroup 9 *cps,* yet both targets were quantified. E: An example of a 2D dPCR plot wherein linkage is observed between *piaB* and serogroup 9 *cps*, high abundance sample.

### Demonstrated Risk of False-Positive Vaccine Serotype Identification in Molecular Testing of URT Samples

To rule out post-PCV circulation of serotype 4 pneumococci (**Fig. 6A**), we assessed linkage between *piaB* and serotype 4 *cps* in saliva samples from PCV10-vaccinated children and their parents. Eighteen of the forty-three (41.9%) tested samples produced quantification for both *piaB* and serotype 4 *cps* in saliva (**Fig. 6C**). That could be interpreted as widespread circulation of serotype 4 *cps* among oral streptococci. Importantly, none of the samples displayed linkage between the targets, indicating that the signal for serotype 4 *cps* originated from a source other than *S. pneumoniae*.

**Figure 6:**
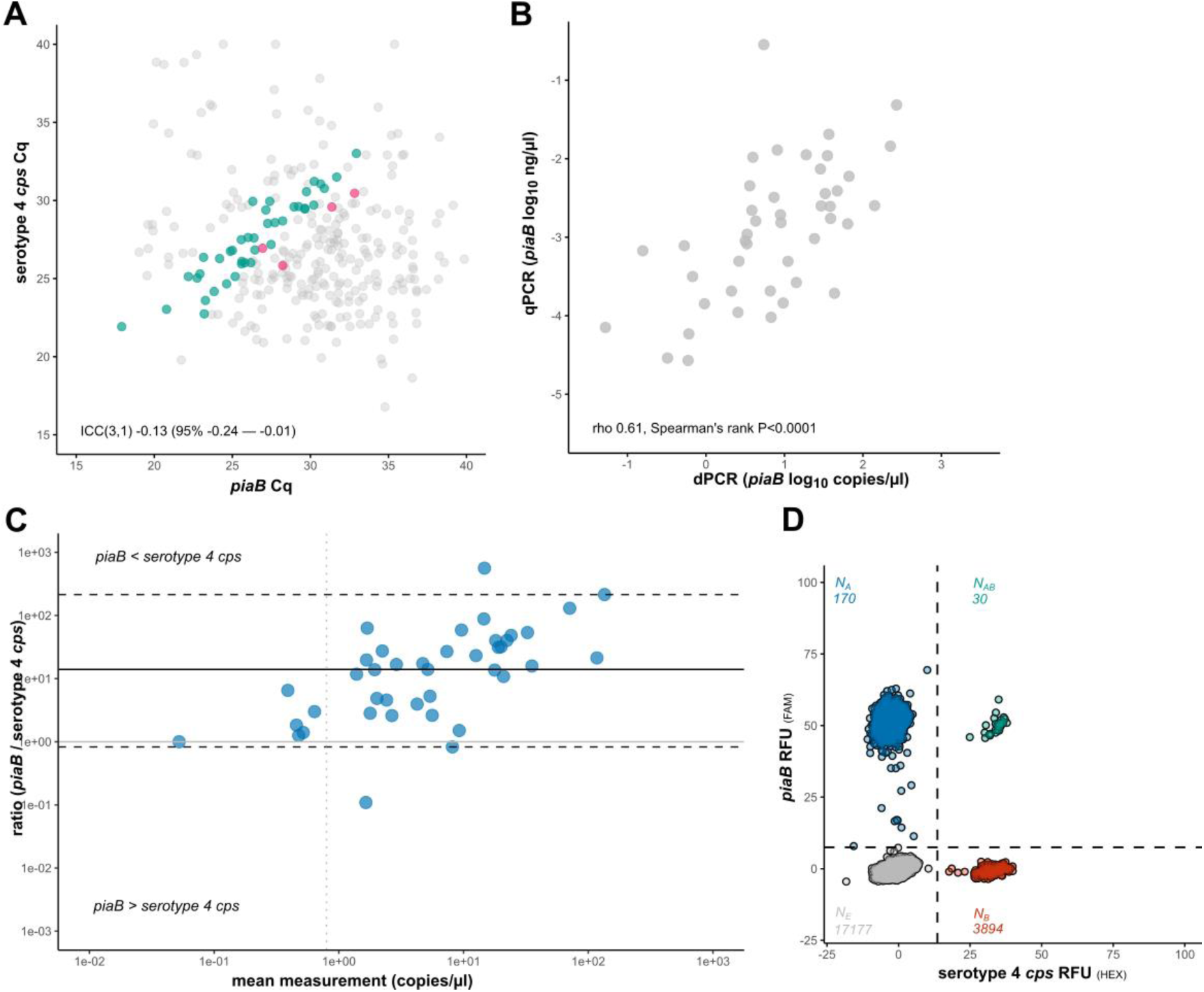
dPCR eliminates false-positive detection of serotype 4, a *cps* that is ubiquitous among oral streptococci. **A:** qPCR-based quantification (in C_q_) of *piaB* and serotype 4 *cps* in DNA extracts of culture-enriched saliva study samples. Samples from children and adults that were selected for dPCR-based linkage analysis are highlighted with green and pink respectively. The remaining samples are colored grey. **B:** The correlation between cultures suggests that pneumococcal presence, as measured by *piaB*, was retained in saliva cultures after preparation of 6-hour cultures (dPCR) from the original overnight cultures (qPCR). **C**: non-parametric Bland-Altman plot of dPCR-based quantification (in copies/µl) *piaB* and serotype 4 *cps* in culture-enriched cell suspensions, samples positive for linkage between *piaB* and serotype 4 *cps* are highlighted with red and negative samples with blue. The vertical line indicates the LoD_95_, dashed horizontal lines the limits of agreement and solid line the bias (median difference between paired *piaB* and serotype 4 *cps* measurements). **D:** An example of a 2D dPCR plot wherein no linkage is observed between *piaB* and serotype 4 *cps.* In this case, the ratio of double-positive to single-positive partitions was not suggestive of linkage (30 × 17177 ≈ 170 × 3894).

## DISCUSSION

Despite abundant data on pneumococcal carriage in children, studies in non-paediatric populations remain limited [8]. Reliance on nasopharyngeal culture has fostered the misconception that pneumococcal colonization is largely absent in adults, despite a relatively high burden of disease in older adults and early-20^th^-century studies documenting substantial adult carriage rates using mouse inoculation methods applied to oral samples [34]. These and other studies suggest that pneumococcal colonization in adults is primarily confined to oral and oropharyngeal mucosal surfaces [9], and that subgroup variation in diagnostic test performance undermines the sensitivity of nasopharyngeal culture [35]. Nonetheless, adult carriage surveys are gaining importance with the rollout of adult vaccination programs.

In highly polymicrobial samples like oral samples, where the relative abundance of the targeted organism is low, culture-based approaches with limited selectivity or electivity often produce discrepancies with culture-independent methods, a longstanding phenomenon known as the ‘great plate count anomaly’ [10]. To yield meaningful data, adult surveys must include oral fluids or oropharyngeal swabs combined with molecular methods, as most carriers would otherwise not be identified [36]. However, lateral gene transfer (LGT) of capsular genes between *S. pneumoniae* and other members of the *Streptococcus mitis* group, common residents of the oral cavity, can compromise the diagnostic specificity of molecular serotyping in airway samples, as observed in several (though not all) epidemiologically relevant assays [5]. Clusters of functionally related genes, such as the capsular operon, are especially prone to LGT, complicating molecular surveillance [37]. To address this, we evaluated digital PCR for its ability to accurately identify *S. pneumoniae* serotypes in highly polymicrobial URT samples, using linkage analysis between *piaB* and serotype-specific targets.

In this study, we adapted the multiplex single intact cell dPCR (‘MuSIC’ dPCR) protocol of McMahon and coworkers for identification and molecular serotyping of *S. pneumoniae* in upper airway specimens [18]. The modified protocol uses a shorter 6-hours incubation to maximize the relative abundance of *S. pneumoniae* intact cells in samples. The results presented in this study show that concordant quantification of *piaB* and *lytA* in culture-enriched saliva correlates with the detection of *S. pneumoniae*, thereby providing support for qPCR-based approaches in prior studies [8]. Furthermore, linkage analysis enabled differentiation between *S. pneumoniae* serogroup 9 *cps* and non-pneumococcal signatures of the same serogroup within individual samples, allowing for accurate molecular serotyping in settings where serotype/serogroup-specific targets circulate in both *S. pneumoniae* and non-pneumococcal subpopulations.

Consistent with previous studies on pathogenic *Escherichia coli* [18, 19], linkage analysis using DNA extracts proved not feasible for physically distant gene targets. In our case, when using purified *S. pneumoniae* DNA the spatial separation between targets far exceeded the typical DNA fragment length of DNA extracts, resulting in minimal *piaB/lytA* linkage percentages (−0.8-0.3%). To overcome this limitation, we performed linkage analysis on intact cells instead, which yielded substantially higher linkage percentages, ranging from 53% to 71%. Given the low levels of *S. pneumoniae* abundance in oral fluids, linkage percentage was estimated independently of abundance [5]. Linkage percentages below the theoretical maximum of 100% were likely to reflect the quantification of extracellular DNA from permeabilized cells, which like DNA extracts, did not enable identification of linkage [19]. In line with this notion, samples from overnight cultures produced low linkage percentages, ranging from 0.9% to 7.6%, which suggested that quantification in these samples was primarily derived from permeabilized cells. We hypothesized that shorter incubation times might yield higher linkage values, based on unpublished findings indicating that *S. pneumoniae* enrichment with gentamicin sheep blood agar predominantly occurs within the first eight hours of incubation. Indeed, a six-hour incubation led to a marked increase in linkage in 4 out of 5 selected study samples. Further tests with suspensions of pneumococci in mid-log-phase indicated that a 6-hour incubation produced maximal linkage compared to longer incubation times. Low percentage of linkage between PCR targets in older cultures was hypothesized to be due to the synthesis of antagonistic compounds during stationary phase of bacterial growth [33], potentially leading to diminished viability and the accumulation of extracellular (and fragmented) DNA. Accordingly, all study samples were thereafter tested for linkage using a 6-hour incubation. Prospective studies aiming to utilize dPCR for linkage analysis should therefore consider the 6-hour cultures over overnight cultures.

Given the possibility of PCR-inhibitory compounds in bacterial suspensions leading to ‘rain’ (partitions with intermediate fluorescence) and false-negative results, we evaluated whether extending the number of PCR cycles could improve the identification of linkage between genes. A modest increase in linkage was observed when the thermocycling program was extended to 55 cycles of annealing and extension, compared to 45 cycles. Contrary to previous reports [31], the addition of lysozyme did not further improve linkage detection. Likewise, the addition of bovine serum albumin did not improve linkage detection (data not shown). Thereafter, the analytical sensitivity of the method was evaluated using probit regression analysis of replicate measurements of serially diluted mid-log-phase *S. pneumoniae* suspensions. A LoD_95_ of 1458 CFU/ml was found, amounting to 0.80 copies/µl.

Accuracy of the method was assessed using mixes of *S. pneumoniae* 9N (*piaB*-positive) with non-pneumococcal serogroup 9 (*piaB-*negative^-^) strains. Linkage between *piaB* and serogroup 9 *cps* was identified provided that the abundance of the *S. pneumoniae* strain was above the LoD95. The linkage value based on serogroup 9 *cps* reflected the ratio of *S. pneumoniae* 9N and non-pneumococcal serogroup 9 strain, whereas the linkage value based on *piaB* solely reflected the *S. pneumoniae* 9N strain.

A set of 43 culture-enriched saliva samples varying in pneumococcal abundance for both *lytA* and *piaB* were tested with dPCR linkage analysis. Prior analysis of these samples with qPCR had suggested positive conditional dependence between *piaB* and *lytA*, as indicated by near-perfect agreement in paired measurements in Bland-Altman analysis [5]. In line with this notion, linkage between *piaB* and *lytA* was observed in culture-enriched saliva provided that quantification of both targets was above the LoD_95_. Linkage was regarded as evidence of positive conditional dependence between *piaB* and *lytA* and the co-occurrence of both targets on a bacterial genome was considered specific to encapsulated *S. pneumoniae* [22]. This finding provided further support for the utility of the Two-to-Tango approach in guiding interpretation of carriage surveillance data on polymicrobial samples. Linkage was also identified between *piaB* and serogroup 6 *cps* assays for samples with concordant quantification of both targets and provided that quantification was above the LoD_95_. In this analysis the *piaB* assay was used as *S. pneumoniae* specific marker and linkage between both assays was regarded as evidence of a serogroup 6 *cps S. pneumoniae*. The serogroup 6 *cps* assay also produced concordant quantification in prior studies [5] and this in conjunction with the findings from dPCR-based linkage analysis, suggested that such assays can be reliably interpreted with qPCR.

Next, the serogroup 9 *cps* assay was chosen as qPCR-based surveillance and nasopharyngeal culture data had suggested simultaneous circulation of *S. pneumoniae* serogroup 9 *cps* and nonpneumococcal *S. mitis* group serogroup 9 *cps.* This assay includes the PCV21 vaccine type (VT) 9N as well as the PCV7/10/13/15/20 VT 9V. Indeed, dPCR analysis demonstrated that the majority of the saliva samples were negative for linkage between *piaB* and serogroup 9 *cps*, a finding suggestive of a non-pneumococcal origin of sequences detected in assay targeting serogroup 9 *cps.* Nevertheless, the saliva samples from four individuals, all of which were positive by nasopharyngeal culture for 9N, displayed linkage between targets ranging from 16% to 58%. These observations highlighted the ability of dPCR-based linkage analysis to control diagnostic specificity and differentiate nonpneumococcal serogroup 9 *cps* signatures from pneumococcal serogroup 9 *cps* signatures. As such, the outlined protocol provides a promising methodology for molecular serotyping in studies utilizing culture-independent techniques. This finding bears particular importance to adult carriage studies where oral fluids and oropharyngeal samples are the sample of choice [8, 9]. It also demonstrates the risk of false-positive pneumococcal serotyping results when certain precautionary measures are not considered (*e.g.,* assessment of concordance). In our previous studies, qPCR results for such assays were altogether excluded from epidemiologic interpretation [5, 13].

This risk was particularly evident for the serotype 4 *cps* assay. Serotype 4 *S. pneumoniae* is a highly invasive serotype and a PCV7-generation VT that continues to cause pneumococcal disease and is at times linked to localized outbreaks [38, 39]. The serotype 4 *cps* PCR assay is in multiple settings associated with widespread circulation of non-pneumococcal *S. mitis* group serotype 4 *cps*. Consequently, molecular serotyping results on oral fluids as well as nasopharyngeal samples, are often positive for serotype 4 *cps* yet negative for *S. pneumoniae*-specific markers (*e.g., piaB* and *lytA*). This poses a risk for studies using culture-independent molecular serotyping, as several reports of high post-PCV serotype 4 carriage may have been confounded by nonpneumococcal *S. mitis* group serotype 4 *cps* [40–42]. By implementing dPCR-based linkage analysis, a simple method of molecular serotyping that does not require DNA extraction, investigators can distinguish *S. pneumoniae* serotype signatures from non-pneumococcal serotype signatures.

Optimized qPCR assays can be easily transferred from qPCR systems to the dPCR platform. On the Qiagen dPCR platform, 26k-well nanoplates can accommodate suspensions of intact bacterial cells, and this platform requires no additional steps, such as droplet generation. In comparison to qPCR, the dynamic range of dPCR is somewhat more limited, with highly concentrated samples requiring prior dilution. To this end, we recommend testing DNA extracts of samples for *S. pneumoniae* with *piaB* and *lytA* by qPCR. Following the identification of *S. pneumoniae* positive samples, these positive samples can be pooled and tested for serotype/serogroup-specific qPCR assays. Individual samples from pools that exhibit serotype/serogroup-specific signal for assays can then be tested individually on the dPCR platform for molecular serotyping (**Fig 6**). Alternatively, *S. pneumoniae* positive samples can be tested directly by dPCR. dPCR technology holds promise for molecular surveillance studies, not only by improving diagnostic specificity but also through its enhanced capability for assay multiplexing [43].

**Fig. 6:**
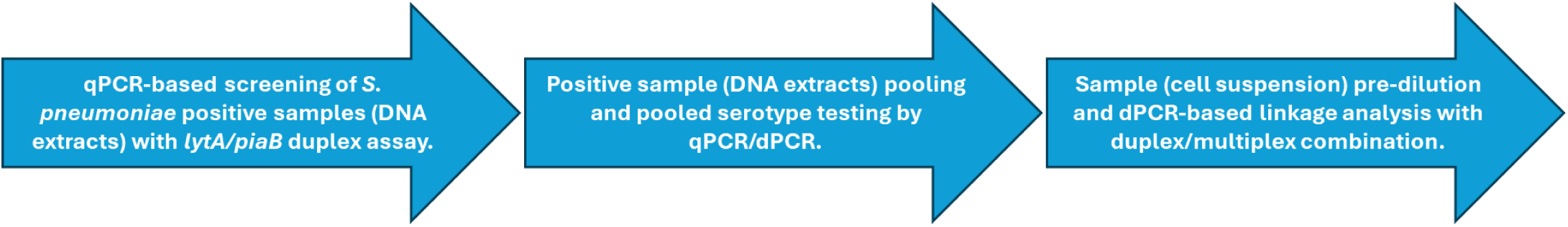
Proposed sample testing workflow. Detection of *S. pneumoniae* in DNA extracts is performed by qPCR, given its lower cost and broad dynamic range. Thereafter, samples classified as *S. pneumoniae* positive are pooled in pools of five samples and tested for a panel of serotype/serogroup-specific assays, either by qPCR or dPCR (*e.g.,* with highly multiplexed dPCR assays). Assays with diagnostic specificity issues can be identified by testing pools of negative samples. Such assays are selected for dPCR-based linkage analysis in duplex with *S. pneumoniae* markers (*piaB*/*lytA*) and harvests of cell suspensions (*i.e.* glycerol supplemented suspensions). Harvests can be diluted according to qPCR quantification from the corresponding DNA extract to avoid exceeding the dPCR upper detection limit.

### Limitations of the study

This study is subject to certain limitations. The protocol requires samples supplemented with glycerol and storage at proprer conditions (*i.e.*, −70°C). Only a limited number of assays were tested with the dPCR-based linkage analysis. In presumably rare cases where a sample harbors multiple *S. pneumoniae* serotypes, including the serotype of interest, alongside an oral streptococcus carrying a homologous serotype signature, false-negative results may arise, especially when the target *S. pneumoniae* serotype is present at low abundance. Such samples are expected to show low linkage percentages between the serotype target and *piaB/lytA*, potentially complicating interpretation.

In conclusion, dPCR-based linkage analysis represents a promising approach for accurate molecular serotyping in polymicrobial upper airways samples. This study demonstrated that dPCR, when applied to intact cells, can reliably detect *S. pneumoniae* in saliva samples from children and adults, and can resolve a *S. pneumoniae* serotype signature from those of non-pneumococcal origin. This is especially important for carriage analyses of adults, since pneumococcal colonization in adults is often confined to polymicrobial sites such as the oral and oropharyngeal mucosa. dPCR has the potential to substantially improve adult carriage surveillance by enabling more accurate molecular detection and serotyping, thereby improving robust evaluation of pneumococcal conjugate vaccine (PCV) impact on carriage.

## STAR★METHODS

Detailed methods are provided in the online version of this paper and include the following:

- KEY RESOURCES TABLE
- RESOURCE AVAILABILITY

- Lead contact
- Materials availability
- Data and code availability
- EXPERIMENTAL MODEL AND STUDY PARTICIPANT DETAILS

- Human subjects
- Microbial strains
- METHOD DETAILS

- Sample collection and laboratory processing
- Study sample culture-enrichment
- Bacterial cell suspensions and CFU assessment
- Nucleic acid extraction
- Primers and probes
- quantitative PCR
- digital PCR
- Baseline correction and RFU thresholding
- Linkage analysis
- Bland-Altman analysis
- Probit regression analysis
- QUANTIFICATION AND STATISTICAL ANALYSIS

- Statistical analysis and data interpretation.

## Data Availability

Data and code availability
All data reported in this paper will be shared by the lead contact upon request.
This paper does not report original code.
Any additional information required to reanalyze the data reported in this paper is available from the lead contact upon request.

## ACKNOWLEDGEMENTS

The authors are very grateful to all the participating children and their families. We thank Spaarne Gasthuis and Streeklab Haarlem for the provision of the original study samples.

## AUTHOR CONTRIBUTIONS

Conceptualization, W.R.M, K.T., and R.M.; methodology, W.R.M, E.H.M.R. van den O., T. N., and J. van V.; investigation, W.R.M.; resources, K.T., and R.M.; data curation, W.R.M, E.H.M.R. van den O., and T. N.; writing – original draft, W.R.M.; review & editing, W.R.M., T. N., E.H.M.R. van den O., J. van V., N.Y.R., K.T., and R.M.; visualization, W.R.M.; supervision, W.R.M., and T. N.; project administration, W.R.M.; funding acquisition, K.T. and R.M.

## RESOURCE AVAILABILITY

### Lead contact

Further information and requests for resources and reagents should be directed to and will be fulfilled by the lead contact: Willem Miellet.

### Materials availability

This study did not generate new unique reagents.

### Data and code availability

- All data reported in this paper will be shared by the lead contact upon request.
- This paper does not report original code.
- Any additional information required to reanalyze the data reported in this paper is available from the lead contact upon request.

## STAR★METHODS

### KEY RESOURCES TABLE

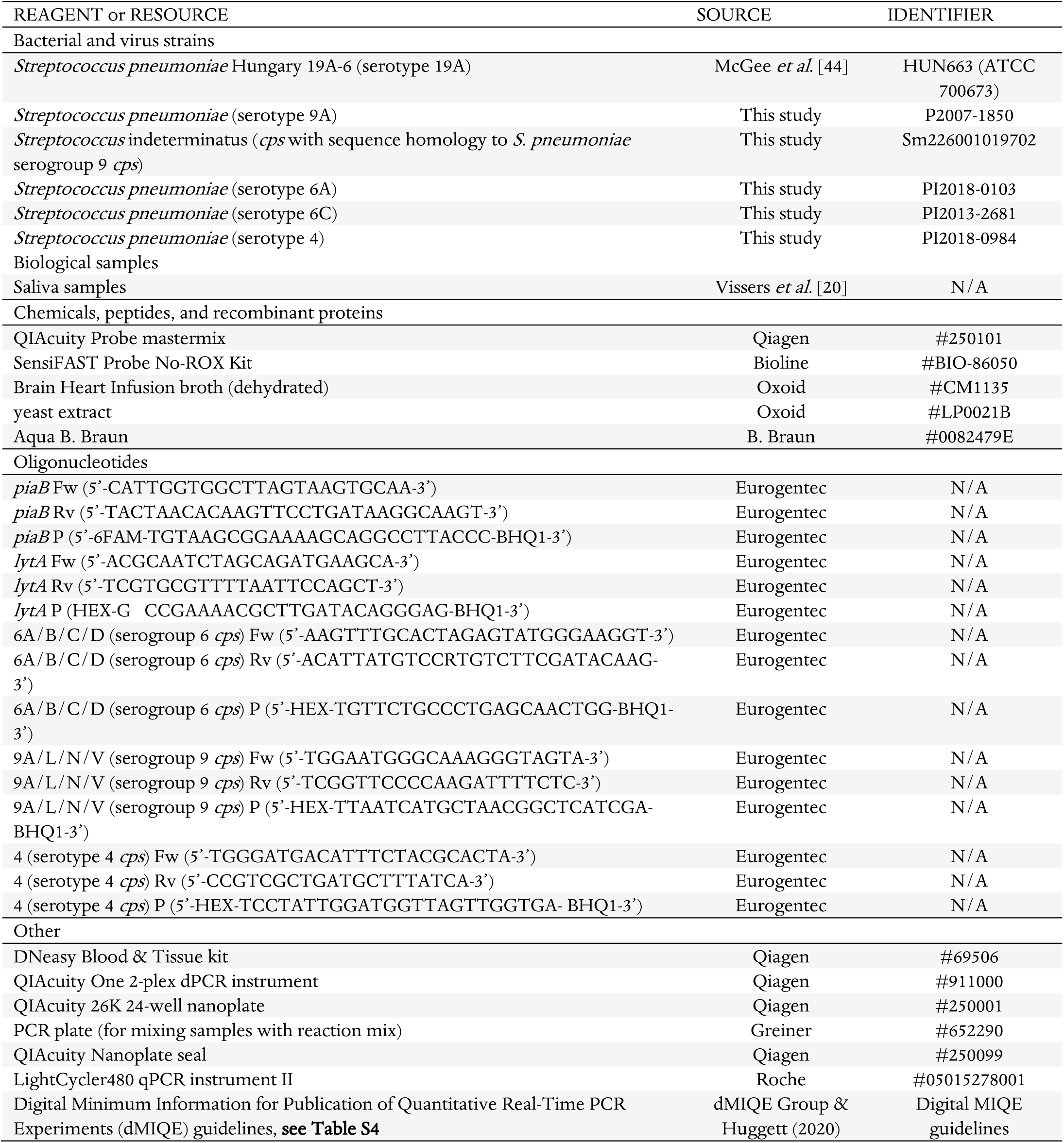

---

**Figure S1:**
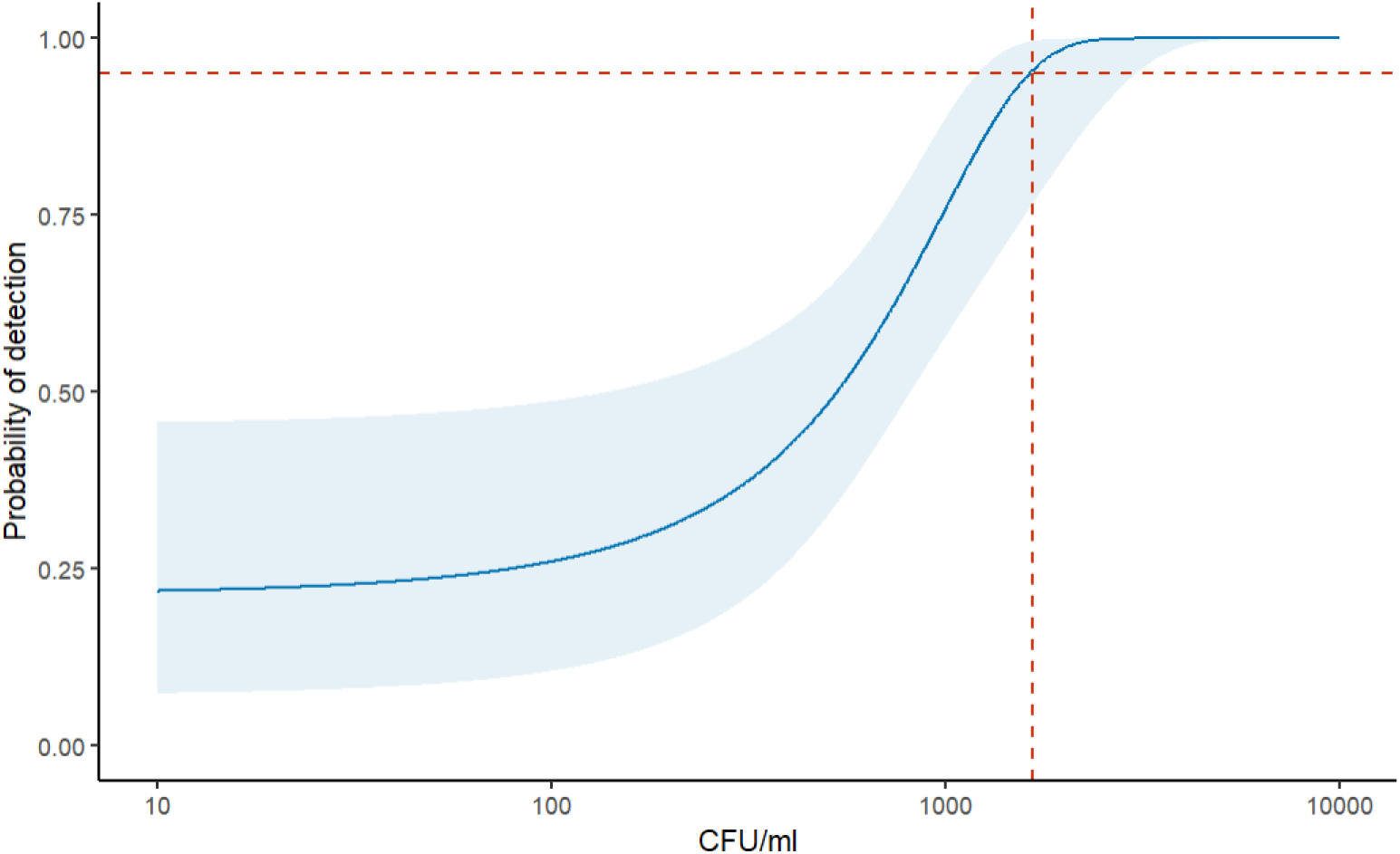
Probit regression analysis of 95% limit of detection (LoD_95_) for the probability of linkage identification between *piaB* and *lytA* given *S. pneumoniae* concentrations (in CFU/ml) in bacterial cell suspensions. The LoD_95_ corresponded to 1664 CFU/ml, which amounts to 9.2 CFU/reaction or 4.4 cp/reaction.

**Figure S2:**
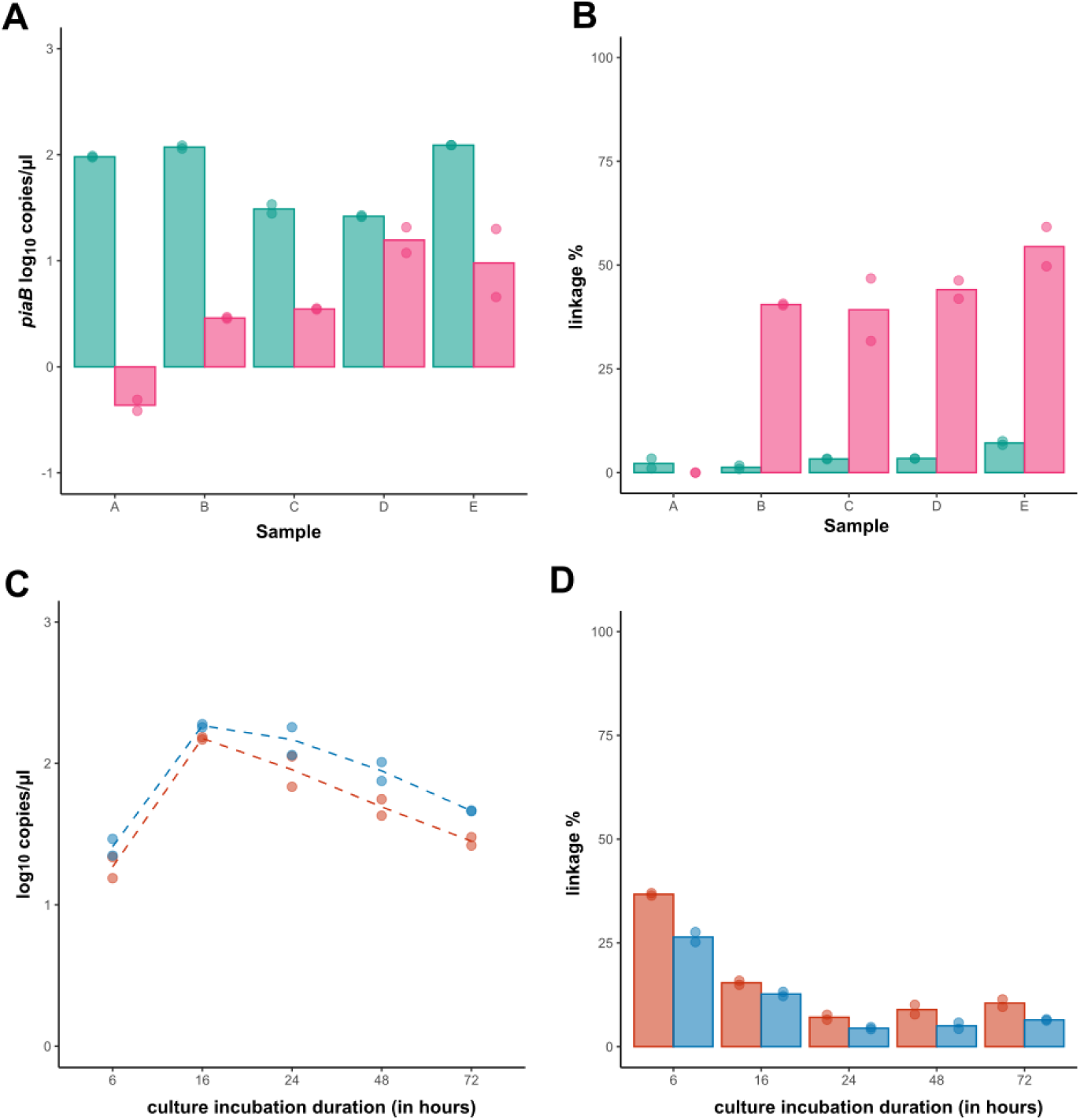
Target quantification and percentage linkage in relation to culture incubation times. **A:** log10-transformed concentrations of *piaB,* with 24-hour (green) cultures and 6-hr (pink) cultures. **B:** Percentage linkage of *piaB* among 24-hour (green) and 6-hour (pink) cultures. **C**: log10-transformed concentrations of *piaB* (red) and *lytA* (blue) at different culture incubation durations (in hours). **D**: Percentage linkage of *piaB* (red) and *lyta* (blue) at different culture incubation durations. Among the observed timepoints, linkage percentage of *piaB* and *lytA* was maximal at 6 hours. Accordingly, 6 hours incubation periods were used for linkage analysis on study samples.

**Table S1:** Strain characteristics.

| Strain name | Catalase | Bile solubility test | Optochin sensitivity | *lytA*<sup>a</sup> | *piaB*<sup>a</sup> | Quellung | *cps*<sup>a</sup> |
| --- | --- | --- | --- | --- | --- | --- | --- |
| Hungary 19A-6 | negative | soluble | sensitive | positive | positive | 19A | serotype 19A |
| P2007-1850 | negative | soluble | sensitive | positive | positive | 9A | serogroup 9 |
| Sm226001019702 | negative | insoluble | resistant | negative | negative | ND | serogroup 9 |
| PI2018-0103 | negative | soluble | sensitive | positive | positive | 6A | serogroup 6 |
| PI2013-2681 | negative | soluble | sensitive | positive | positive | 6C | serogroup 6 |
| PI2018-0984 | negative | soluble | sensitive | positive | positive | 4 | serotype 4 |

a: Tested by qPCR; ND: not determined.

**Table S2:**
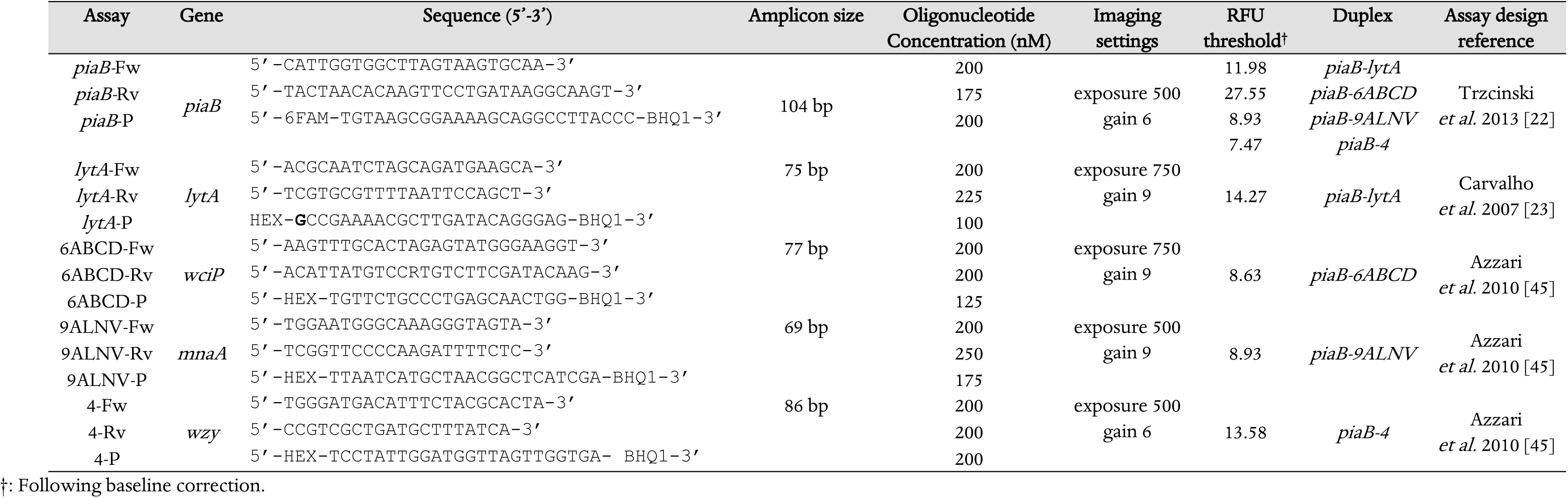
digital PCR primer and probes.

## REFERENCES

1. Ganaie FA, Beall BW, Yu J, et al. Update on the evolving landscape of pneumococcal capsule types: new discoveries and way forward. Clinical Microbiology Reviews 2025: e00175–24.

2. Dagan R, Muallem M, Melamed R, Leroy O, Yagupsky P. Reduction of pneumococcal nasopharyngeal carriage in early infancy after immunization with tetravalent pneumococcal vaccines conjugated to either tetanus toxoid or diphtheria toxoid. Pediatr Infect Dis J 1997; 16(11): 1060–4.

3. Auranen K, Rinta-Kokko H, Goldblatt D, et al. Colonisation endpoints in Streptococcus pneumoniae vaccine trials. Vaccine 2013; 32(1): 153–8.

4. Hendley JO, Sande MA, Stewart PM, Gwaltney JM, Jr. Spread of Streptococcus pneumoniae in families. I. Carriage rates and distribution of types. J Infect Dis 1975; 132(1): 55–61.

5. Miellet WR, van Veldhuizen J, Litt D, et al. A spitting image: molecular diagnostics applied to saliva enhance detection of Streptococcus pneumoniae and pneumococcal serotype carriage. Frontiers in Microbiology 2023; 14.

6. Wyllie AL, Rots NY, Wijmenga-Monsuur AJ, van Houten MA, Sanders EAM, Trzcinski K. Saliva as an alternative sample type for detection of pneumococcal carriage in young children. Microbiology (Reading) 2023; 169(10).

7. Wrobel-Pawelczyk I, Ronkiewicz P, Wanke-Rytt M, et al. Pneumococcal carriage in unvaccinated children at the time of vaccine implementation into the national immunization program in Poland. Sci Rep 2022; 12(1): 5858.

8. Miellet WR, Almeida ST, Trzciński K, Sá-Leão R. Streptococcus pneumoniae carriage studies in adults: Importance, challenges, and key issues to consider when using quantitative PCR-based approaches. Frontiers in Microbiology 2023; 14.

9. Miellet WR, Mariman R, van Veldhuizen J, et al. Impact of age on pneumococcal colonization of the nasopharynx and oral cavity: an ecological perspective. ISME communications 2024; 4(1): ycae002.

10. Staley JT, Konopka A. Measurement of in situ activities of nonphotosynthetic microorganisms in aquatic and terrestrial habitats. Annu Rev Microbiol 1985; 39: 321–46.

11. McLean AR, Torres-Morales J, Dewhirst FE, Borisy GG, Mark Welch JL. Site-tropism of streptococci in the oral microbiome. Molecular oral microbiology 2022; 37(6): 229–43.

12. Velsko IM, Perez MS, Richards VP. Resolving Phylogenetic Relationships for Streptococcus mitis and Streptococcus oralis through Core-and Pan-Genome Analyses. Genome Biol Evol 2019; 11(4): 1077–87.

13. Miellet WR, van Veldhuizen J, Litt D, et al. It Takes Two to Tango: Combining Conventional Culture With Molecular Diagnostics Enhances Accuracy of Streptococcus pneumoniae Detection and Pneumococcal Serogroup/Serotype Determination in Carriage. Front Microbiol 2022; 13: 859736.

14. Parker AM, Jackson N, Awasthi S, et al. Upper respiratory Streptococcus pneumoniae colonization among working-age adults with prevalent exposure to overcrowding. Microbiology Spectrum 2024; 12(8): e00879– 24.

15. Almeida ST, Paulo AC, Simoes AS, Ferreira B, Sa-Leao R. Streptococcus pneumoniae carriage in adults during the COVID-19 pandemic in Portugal: dominance of serotypes included in broader PCVs and of serotype 3. mSphere 2025: e0008225.

16. Boelsen LK, Dunne EM, Gould KA, et al. The Challenges of Using Oropharyngeal Samples To Measure Pneumococcal Carriage in Adults. mSphere 2020; 5(4).

17. Satzke C, Turner P, Virolainen-Julkunen A, et al. Standard method for detecting upper respiratory carriage of Streptococcus pneumoniae: updated recommendations from the World Health Organization Pneumococcal Carriage Working Group. Vaccine 2013; 32(1): 165–79.

18. McMahon TC, Blais BW, Wong A, Carrillo CD. Multiplexed Single Intact Cell Droplet Digital PCR (MuSIC ddPCR) Method for Specific Detection of Enterohemorrhagic E. coli (EHEC) in Food Enrichment Cultures. Front Microbiol 2017; 8: 332.

19. He L, Simpson DJ, Ganzle MG. Detection of enterohaemorrhagic Escherichia coli in food by droplet digital PCR to detect simultaneous virulence factors in a single genome. Food Microbiol 2020; 90: 103466.

20. Vissers M, Wijmenga-Monsuur AJ, Knol MJ, et al. Increased carriage of non-vaccine serotypes with low invasive disease potential four years after switching to the 10-valent pneumococcal conjugate vaccine in The Netherlands. PLoS One 2018; 13(3): e0194823.

21. Krone CL, Wyllie AL, van Beek J, et al. Carriage of Streptococcus pneumoniae in aged adults with influenza-like-illness. PLoS One 2015; 10(3): e0119875.

22. Trzcinski K, Bogaert D, Wyllie A, et al. Superiority of trans-oral over trans-nasal sampling in detecting Streptococcus pneumoniae colonization in adults. PLoS One 2013; 8(3): e60520.

23. Carvalho Mda G, Tondella ML, McCaustland K, et al. Evaluation and improvement of real-time PCR assays targeting lytA, ply, and psaA genes for detection of pneumococcal DNA. J Clin Microbiol 2007; 45(8): 2460–6.

24. Trypsteen W, Vynck M, De Neve J, et al. ddpcRquant: threshold determination for single channel droplet digital PCR experiments. Analytical and bioanalytical chemistry 2015; 407: 5827–34.

25. Huggett JF. The digital MIQE guidelines update: minimum information for publication of quantitative digital PCR experiments for 2020. Clinical chemistry 2020; 66(8): 1012–29.

26. Regan JF, Kamitaki N, Legler T, et al. A rapid molecular approach for chromosomal phasing. PloS one 2015; 10(3): e0118270.

27. Altman DG, Bland JM. Interaction revisited: the difference between two estimates. BMJ 2003; 326(7382): 219.

28. Bland JM, Altman DG. Measuring agreement in method comparison studies. Stat Methods Med Res 1999; 8(2): 135–60.

29. Ahlers FM, Litt DJ, Jansen van Rensburg MJ, et al. Utilizing large and diverse bacterial genome datasets to improve the detection and identification of Streptococcus pneumoniae via PCR-based diagnostics. Microb Genom 2025; 11(6).

30. Lei S, Gu X, Zhong Q, Duan L, Zhou A. Absolute quantification of Vibrio parahaemolyticus by multiplex droplet digital PCR for simultaneous detection of tlh, tdh and ureR based on single intact cell. Food control 2020; 114: 107207.

31. Diebold PJ, New FN, Hovan M, Satlin MJ, Brito IL. Linking plasmid-based beta-lactamases to their bacterial hosts using single-cell fusion PCR. Elife 2021; 10.

32. Wang MY, Olson BH, Chang JS. Improving PCR and qPCR detection of hydrogenase A (hydA) associated with Clostridia in pure cultures and environmental sludges using bovine serum albumin. Appl Microbiol Biotechnol 2007; 77(3): 645–56.

33. Mellroth P, Daniels R, Eberhardt A, et al. LytA, major autolysin of Streptococcus pneumoniae, requires access to nascent peptidoglycan. Journal of Biological Chemistry 2012; 287(14): 11018–29.

34. Krone CL, van de Groep K, Trzcinski K, Sanders EA, Bogaert D. Immunosenescence and pneumococcal disease: an imbalance in host-pathogen interactions. Lancet Respir Med 2014; 2(2): 141–53.

35. Mulherin SA, Miller WC. Spectrum bias or spectrum effect? Subgroup variation in diagnostic test evaluation. Ann Intern Med 2002; 137(7): 598–602.

36. Austrian R. The pneumococcus at the millennium: not down, not out. The Journal of infectious diseases 1999: S338–S41.

37. Novick A, Doolittle WF. Horizontal persistence and the complexity hypothesis. Biology & Philosophy 2019; 35(1): 2.

38. Kellner JD, Ricketson LJ, Demczuk WHB, et al. Whole-Genome Analysis of Streptococcus pneumoniae Serotype 4 Causing Outbreak of Invasive Pneumococcal Disease, Alberta, Canada. Emerg Infect Dis 2021; 27(7): 1867– 75.

39. Gladstone RA, Siira L, Brynildsrud OB, et al. International links between Streptococcus pneumoniae vaccine serotype 4 sequence type (ST) 801 in Northern European shipyard outbreaks of invasive pneumococcal disease. Vaccine 2022; 40(7): 1054–60.

40. Amodio E, Tramuto F, De Francisci V, et al. Pneumococcal carriage in a large Sicilian sample population: impact on the current epidemiological scenario and implications for future vaccination strategies. Front Cell Infect Microbiol 2024; 14: 1467320.

41. Ricketson LJ, Lidder R, Thorington R, et al. PCR and Culture Analysis of Streptococcus pneumoniae Nasopharyngeal Carriage in Healthy Children. Microorganisms 2021; 9(10).

42. Warda K, Amari S, Boureddane M, et al. Changes in pneumococcal serotypes distribution and penicillin resistance in healthy children five years after generalization of PCV10. Heliyon 2024; 10(4): e25741.

43. Whale AS, Huggett JF, Tzonev S. Fundamentals of multiplexing with digital PCR. Biomol Detect Quantif 2016; 10: 15–23.

44. McGee L, McDougal L, Zhou J, et al. Nomenclature of major antimicrobial-resistant clones of Streptococcus pneumoniae defined by the pneumococcal molecular epidemiology network. J Clin Microbiol 2001; 39(7): 2565–71.

45. Azzari C, Moriondo M, Indolfi G, et al. Realtime PCR is more sensitive than multiplex PCR for diagnosis and serotyping in children with culture negative pneumococcal invasive disease. PLoS One 2010; 5(2): e9282.

